# External validation of a clinical decision rule and neuroimaging rule-out strategy for exclusion of subarachnoid haemorrhage in the emergency department: A prospective observational cohort study

**DOI:** 10.1101/2021.05.26.21257212

**Authors:** Tom Roberts, Robert Hirst, William Hulme, Daniel Horner, on Behalf of TERN

**Author notes:** Corresponding Author: Dr Tom Roberts, 07894 234121, 12 Hamilton Road, Bristol, BS3 1PB.

## Abstract

**Background:** The investigation of suspected subarachnoid haemorrhage (SAH) presents a diagnostic dilemma. The limited sensitivity of a negative CT brain scan has historically mandated hospital admission and a lumbar puncture to look for evidence of blood in the cerebrospinal fluid. However, emerging evidence has suggested the sensitivity of clinical decision rules and modern CT imaging protocols within early onset of symptoms, may be sufficient to exclude the diagnosis.

**Methods:** A prospective, multi-centre, observational study of consecutive adult patients with acute severe non-traumatic headache presenting to emergency departments. We plan to recruit 9000 patients from over a hundred sites across the UK. The primary outcome is adjudicated SAH as defined by neuroimaging or cerebrospinal fluid findings consistent with the diagnosis.

Data will be collected on clinical history, examination findings, phlebotomy and imaging results. All participants will be followed for 28-days to identify SAH and other clinically relevant outcomes using case note review, and later Hospital Episode Statistics. A proportionate opt-out model of consent will be used to maximise patient recruitment and study generalisability.

**Discussion:** Whilst there is increasing evidence that early neuroimaging strategies for the diagnosis of SAH are very sensitive, there have been no large studies to confirm this in the UK population. Furthermore, the test characteristics of CT brain beyond 6 hours from onset are not well understood and there is limited biological plausibility for this defined time cutpoint. Finally, the performance of the Ottawa clinical decision rule has shown promise in the Canadian population. However, its performance in the UK has not been studied and there are concerns that due to the low specificity it may result in increased, rather than decreased rate of investigations. This study will therefore aim to assess the test characteristics of both a CT brain up to 24h from presentation and the Ottawa SAH clinical decision rule.

## Background

Acute headache accounts for between 1 – 2% of all Emergency Department (ED) attendances, estimated at >350,000 United Kingdom (UK) patients per year. [1–3] A significant proportion of these patients will have serious pathology as the underlying cause of headache. This figure has been as high as 10% in some studies. [4] Unfortunately, subjective clinical headache features and examination findings are often unreliable discriminators. [2] Emergency clinicians pursue high rates of invasive investigation and hospital admissions in order to avoid diagnostic error and reassure patients.

Such defensive strategies are not without harm. Advanced neuroimaging can lead to incidental findings in over 2% of patients, which can create anxiety and necessitate further testing or surgical procedures. [5] In addition, lumbar puncture to definitively exclude subarachnoid haemorrhage (SAH) can prolong hospital stay and lead to recognised complications. [6] Recent evidence also suggests the benefits of this additional invasive test only apply to a small proportion of Computerised Tomography (CT) negative patients with a high pre-test probability. [7] High rates of investigation in headache patients can also lead to significant opportunity cost and reinforce ED attendance patterns.

In a large emergency medicine case series, over 50% of headaches were classed as primary on final diagnosis. [2] There is also increasing guidance from the National Institute for Health and Care Excellence (NICE) regarding the avoidance of acute neuroimaging when feasible. [8] Emergency clinicians are faced with the challenge of selecting appropriate patients to undergo invasive testing, without missing significant underlying causes, in the absence of validated risk prediction.

Several studies have recently focussed on the sensitivity of unenhanced CT brain imaging in isolation, for the exclusion of SAH in patients presenting early to secondary care. Perry et al. first reported a high sensitivity for CT brain imaging if performed within 6 hours in 2011. [9] The described sensitivity of 100% (95% CI, 97%–100%) was later supported by a systematic review and meta-analysis including over 8000 patients in 2016. This study reported a pooled sensitivity of 98.7% (95% CI 97.1% to 99.4%) when CT brain was performed within 6 hours of headache onset in patients without focal neurological deficit. [10] Although compelling, these results are yet to be adopted into national consensus guidelines or routine clinical practice, due to limited validation studies and some authors disputing the high reported sensitivity. [11,12] Indeed, recent implementation work by Perry demonstrated a lower sensitivity of 95.5% (95% CI 89.8% to 98.5%). [13]

Several strategies have recently been published to aid clinicians in the selection of headache patients who may not require any CT imaging. Clinical decision rules are well established in emergency medicine practice as unidirectional decision aids that can support the omission of advanced imaging and limit invasive testing. Such rules have been well received and adopted for head injury, neck injury and chest imaging in trauma. [14–16] Similar rules have been derived to limit invasive testing in acute headache [17,18] and a recent prospective validation of the Ottawa SAH clinical decision rule demonstrated a sensitivity of 100% (95% CI 98.1% to 100%). [13] These rules are yet to be validated in a European population.

UK validation of a 6-hour CT brain only rule out strategy for exclusion of acute SAH in ED patients presenting with headache was recently listed within the top 15 emergency medicine research priorities by patients and clinicians. [19] Further, the accuracy of CT brain in diagnosing SAH at 12 and 24 hours has recently been identified as a research priority by NICE. [20] Use of a highly sensitive decision rule to predict the need for invasive testing could further limit neuroimaging, lumbar puncture and invasive testing within the acute sector.

## Methods and Design

### Study objectives

#### Primary Objective

1. To externally validate a 6-hour CT brain rule out strategy for alert (defined as awake and fully orientated or GCS 15/15) patients presenting with acute non-traumatic headache, where there is clinical concern for SAH.

#### Secondary Objectives

1. To understand the change in sensitivity of CT brain imaging in patients presenting with acute severe headache over hourly intervals from 6 to 24 hours after headache onset.
2. To validate the Ottawa subarachnoid haemorrhage clinical decision rule in patients presenting with acute non-traumatic headache.
3. To report the prevalence of subarachnoid haemorrhage in patients under 40 years of age

### Study Design and Conduct

Consecutive patients presenting to EDs or equivalent secondary care services with non-traumatic headache reaching maximal intensity within 1 hour, will be screened and selected by trained doctors or research nurses for inclusion. It is recognised that historically the majority of these patients would have accessed secondary care via the ED. However, with the advent of Same Day Emergency Care (SDEC) and the growth of ambulatory medicine within the UK, in addition to the impact of the novel coronavirus (COVID-19), some sites may triage patients with acute severe headache to other services within the acute hospital environment. [21] The services are defined as ‘equivalent secondary care services’ in the context of this study.

Essential clinical headache features will be collected prospectively at screening using dedicated inclusion checklists. All other demographic, historical and clinical examination data will be collected at the point of clinical review by the care team and noted in patient care records as standard. Relevant data will be subsequently extracted from patient records by the care team and transferred to anonymised case report forms.

Patients will be identified and essential data collected prospectively but, depending on site resources, some data may be collected retrospectively. Follow up will occur through the clinical care team (including research nurse support) at 24 – 72 hours to ascertain results of neuroimaging, additional investigations, hospital admission and initial diagnostic coding. Further follow up will occur at 28 days by research teams through case note review and primary care contact, to determine any episodes of reattendance, final diagnosis and clinical outcome.

### Participants

#### Inclusion Criteria

- Age 18 years or older
- Presenting with non-traumatic acute headache that reaches maximal intensity within one hour to UK EDs or equivalent acute secondary care services (including Ambulatory Care)
- Alert (defined as awake and fully orientated or GCS 15/15)

#### Exclusion Criteria

- Direct head trauma in the previous 7 days.
- Returning for reassessment of the same headache within the recruitment period.
- Established diagnosis of SAH, brain neoplasm, ventricular shunt or hydrocephalus prior to attendance at the ED.
- Focal neurological deficit.
- Headache with onset >14 days prior to attendance.
- Recurrent headaches (three or more headaches of similar character and intensity as presenting headache).
- Transfer from another hospital with confirmed SAH.
- Prisoner presenting to ED or secondary care.
- Patient currently detained under the Mental Health Act presenting to ED or secondary care.

### Outcome Measures

There is no universally agreed definition for SAH. The following criteria will therefore be used to inform adjudication of a positive case.

#### Any of

- I) Subarachnoid blood present on unenhanced CT brain* reported by a trained radiologist.
- II) Subarachnoid blood present on CT-Angiogram or MR-Angiogram reported by a trained radiologist.
- IIIa) CSF findings consistent with SAH according to the 2008 National Biochemist reporting guideline (outlined in table 1). [22,23]

**Table 1.** Possible Outcomes of Lumbar Puncture.

| Outcome | NBA, AU | NOA |
| --- | --- | --- |
| Positive for blood in CSF | >0.007 | 0 |
|  | >0.007 | ≤0.1 |
|  | >0.007 | >0.1 |
| Negative for blood in CSF | ≤0.007 | 0 |
|  | ≤0.007 | <0.1 |
| Inconclusive | ≤0.007 | ≥0.1 |
| CSF sample uninterpretable; unsuitable for analysis, e.g. insufficient sample, exposed to light, blood contamination. Methemoglobin to be treated as oxyhemoglobin.<br>AU = absorbance units; CSF = cerebrospinal fluid; NBA = net bilirubin absorbance; NOA = net oxyhemoglobin absorbance. |  |  |

The vast majority of UK laboratories processing CSF samples adhere to the 2008 clinical biochemistry guidelines. However, for those sites identified as using xanthochromia or red blood cells in lieu of 2008 criteria, the study team will adopt criteria stipulated previously by Perry et al [9]:

- IIIb) Visible xanthochromia
- IIIc) Red blood cells (>5×10^6^/L) in the final tube of cerebrospinal fluid collected **<u>and</u>** an aneurysm identified on cerebral angiography (digital subtraction, computed tomography, or magnetic resonance angiography)

*Site CT characteristics will be collected to include minimum criteria applied for inclusion in the final analysis, as follows:

○ A 3^rd^ generation multi-slice scanner (4 to 320 slices/rotation).
○ 5-7.5mm cuts for brain.
○ 2.5 – 5mm cuts for the posterior fossa.

In addition to the above objective markers, all potential diagnoses of SAH will be independently adjudicated by a separate committee of clinicians, including a neurosurgeon, an emergency physician and a neuro-intensivist blinded to the impression of treating clinicians. Due to the potential for cases of traumatic LP and incidental aneurysms to be mis-diagnosed as a SAH using criteria IIIc, the adjudication committee will have the final rule on case definition. Any decisions will be documented in writing with full minutes of the discussions leading to the decisions stored alongside study data.

We will ensure all cases of SAH within the cohort are captured by performing follow up to 28 days from presentation. Case note review and primary care contact will be integrated to determine reference standard diagnosis, clinical outcomes and mortality. We will also pursue HES data using the 4-digit diagnostic codes for subarachnoid haemorrhage following database lock.

Radiologists reporting all imaging modalities will be provided with clinical information via their normal local requesting procedures and report according to local policy. CT scans reported overnight will be included in the study. The grade of radiologist, use of telemedicine reporting and verified consultant reports will be included in the dataset. The final verified consultant report will be utilised when adjudicating the diagnoses of SAH.

### Consent

Research in an emergency setting is always challenging due to high clinical work load and the proportion of eligible patients presenting out of hours. Clinicians have additional levels of service demand during night shifts and research teams are often not available to support recruitment. However, it is vital that patients presenting out of hours with acute severe headache are offered the opportunity to participate in this research, and it is also vital to the integrity of the research methodology that these patients are included within a consecutive sample, to ensure the findings are generalizable.

To minimize the risk of recruitment bias in our sample, this study plans to recruit 24 hours a day, 7 days a week, at all sites. To achieve this, there will be extensive departmental awareness of the study raised at each site, and we will use trained clinicians to recruit to the study as part of routine care, in addition to research teams. As such, it is essential that we minimize the administrative burden for these busy clinicians. We have chosen an opt-out consent process to facilitate this, designed to ensure patients are robustly informed and provided with tiered opportunity to ask questions/engage with the research at their preference. This method will minimize the risk of missing a significant cohort of patients presenting overnight, minimising the burden on clinical staff recruiting during standard working hours but maximising the generalisability of the research.

This study involves no change to clinical care and no additional interventions for participants. It therefore carries no clinical risk. It is well recognised that patients attending the ED or secondary care with acute headache are in pain and at a time of acute anxiety/distress and often require strong and sedating analgesia. As such, it will be impractical and potentially distressing to seek fully informed consent in an emergency setting. We will therefore adopt a proportionate approach using an opt-out strategy, as supported by the Health Research Authority (HRA). [24]

We will display relevant materials in the appropriate areas of every participating ED or secondary care environment, describing the study and providing assurance that clinical care will not be affected in any way. We will offer individual patient information sheets (PIS) at the point of clinical assessment, with a description of the study and identified point of site contact for every patient enrolled to the study. Staff in the ED or secondary care environment will be available at initial clinical assessment and on request, to speak to any participant or their next of kin. All PIS documents will be numbered and linked to the study inclusion checklist. Completion of this checklist will ensure a robust process for ensuring all eligible patients have received the opt-out information. In addition, the PIS will contain links for headache support groups and patient information.

All patients who wish to opt-out will be highlighted on a specific opt-out log. The opt-out log will be cross-referenced with the inclusion checklists and REDCap database at 3 different time points, to ensure that any patient expressing a wish to opt out of the study has been acknowledged and no further data recorded:

1. Prior to initial data entry
2. Retrospectively by the study team at the end of each week
3. 4 weeks after data-collection has finished

Any patients deemed eligible for the study but not present on the opt-out log will be included in the study and data transcribed to case report forms as per study procedures. Sites will be offered a tiered strategy for further/additional patient information, including a PIS to be mailed to a home address or research nurse contact (face to face if in hospital, or by telephone).

This approach, targeted to the individual, is considered to constitute active recruitment as per paragraph 21 of the NIHR Clinical Research Network (CRN) recruitment policy document. [25] In addition, this methodology for observational research has been used with ethical approval and CRN portfolio adoption/accrual for multiple ED projects, including the PAT-POPS study, the AHEAD study and most recently the HEAD study [IRAS 258618, REC 19/SW/0089, CPMS 41849]. [26,27] The opt-out consent process is outline in Figure 1.

**Figure 1.**
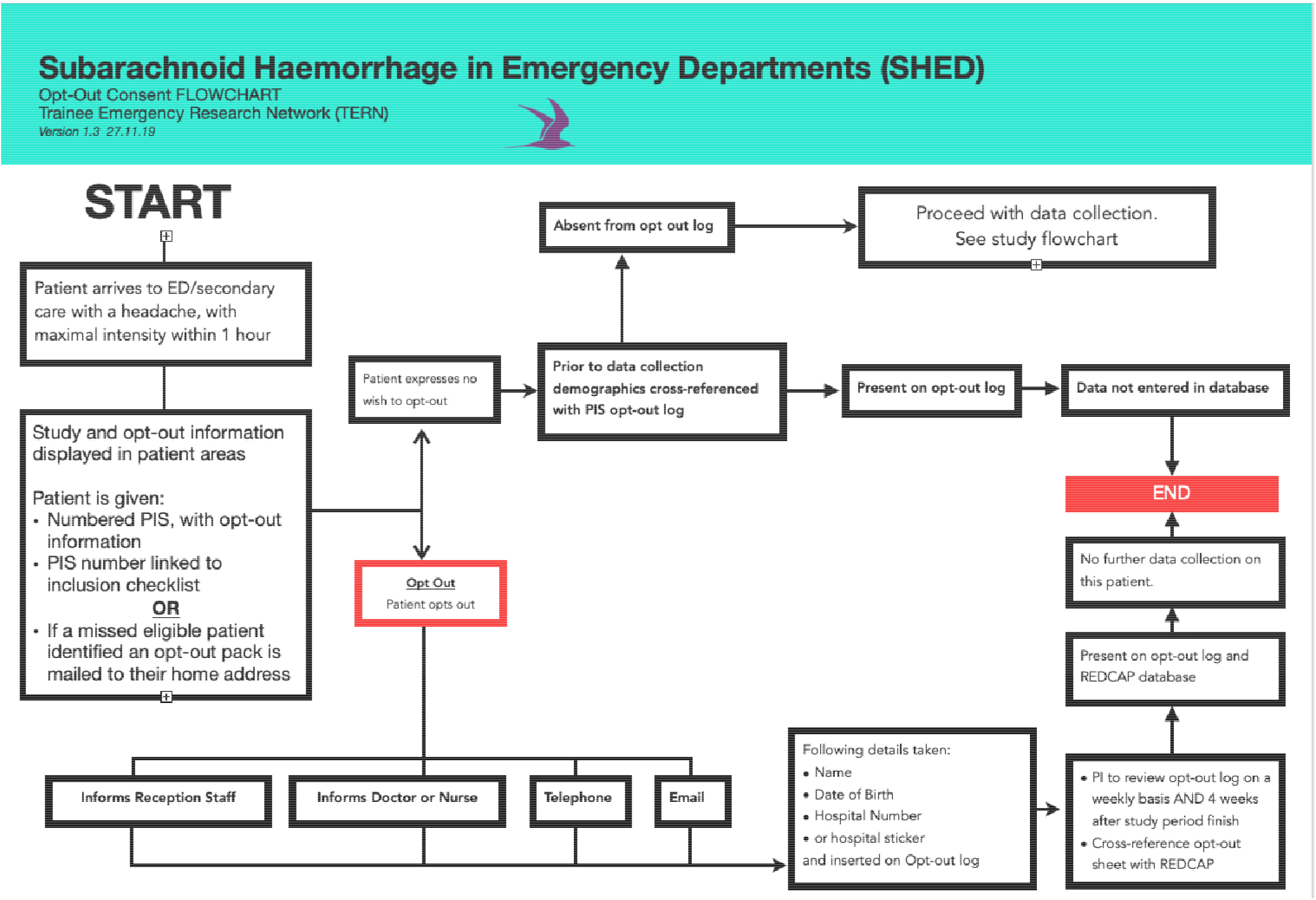
Opt-out consent process

### Withdrawal

Participants will be allowed to withdraw from the study at any point. We will not collect any further data from these participants; due to the opt-out methodology, any patient who expresses a wish to withdraw from the study will have any collected data deleted.

### Administration

All outcome measures will be recorded on an electronic questionnaire and case report forms, REDCap. REDCap is fully compliant with 21 CFR Part 11, GDPR, ISO 27001 and ISO 9001. [28,29] REDCap will be password protected and only accessible to certain members of the study team. Patient identifiable data will never be downloaded or transferred along with research data. Patient identifiable data will be restricted only to the Chief Investigator, be password protected, encrypted and be stored on a secure database as outlined above. Research data will be downloaded once only for the purpose of statistical analysis. The data will be downloaded onto a secure, password protected, University computer system.

### Data Storage

Data will be stored electronically for 5 years by the University Hospital of Bristol and Weston NHS Foundation Trust.

### Patient and Public Involvement

There has been no direct PPI in the design of this study. However, a prospective evaluation of a CT brain scan rule-out pathway without recourse to lumbar puncture in patients with suspected SAH was listed in the top 15 research priorities for Emergency Medicine in 2017 which incorporated both patient and public involvement. [19]

## Statistical Analysis Plan

### Sample size calculation

Given our primary hypothesis is to assess the sensitivity of 6-hour CT brain rule out strategies within our cohort, we have performed sample size calculations designed to power appropriately for this outcome. Prevalence figures for acute SAH in alert patients with atraumatic headache attending the emergency department range from 2% to 7.5%. We will assume a prevalence of 5%. [7,9,10,13]

Prior Canadian work suggests emergency physicians would accept a SAH rule with a sensitivity of 99%, i.e., the rule missing no more than 1 in 100 SAHs is acceptable. [30] For our study we have extrapolated that a similar level of sensitivity would be required for CT Brain imaging in the same cohort of patients. We choose the minimum sample size to ensure we can detect a sensitivity whose 95% confidence interval is above and does not include 98% with at most 3 missed cases (and where 4 or more misses indicates failure to validate the Ottawa SAH rule).

We will use exact (Pearson-Clopper) binomial confidence limits for the sensitivity as the normal approximation fails when the sensitivity is near 100%. The alpha-level is 5% (i.e., we will use 95% confidence intervals). To ensure a lower 95% confidence limit of above 98% with 3 or fewer SAH misses, 436 positive SAH cases are required. The observed sensitivity for 3 missed cases in 436 is 99.3%. With an estimated prevalence of 5%, this requires a total sample size of 436/0.05 = 8,720 attendances for atraumatic headache. To protect against unexpectedly low SAH prevalence we aim to collect n=9000 attendances over the duration of the data collection period.

Using previous data from prospective observational studies, we would expect approximately 30% of patients with suspected subarachnoid haemorrhage to undergo CT imaging within 6 hours. [13] As such, this large sample size should provide a nested cohort of individuals undergoing early CT; we will use data from these patients to assess the diagnostic tests characteristics associated with an early CT only rule out strategy at various timepoints.

Our primary objective is to externally validate the proposed 6-hour CT brain rule out strategy for alert (defined as awake and fully orientated or GCS 15/15) patients presenting with acute non-traumatic headache with maximal intensity within 1 hour. Due to new evidence, those patients with a documented haemoglobin of less than 10g/L will be excluded from the primary analysis but will be included in all secondary analysis. [13,31]

We will use funnel plots to assess for site variation and inconsistency between centres across various stages of the project.

### Recruitment timeline

Current evidence suggests atraumatic headache accounts for between 1 – 2% of all ED attendances. In an average teaching hospital or foundation trust with an attendance of 100,000 per annum, this equates to 125 patients per month attending with headache. It is currently unknown how many of these patients present with headache that reaches maximal intensity within one hour. We have therefore conservatively estimated that 25% of patients presenting with headache will reach maximal intensity within one hour, per month; this would provide 31 patients per month for recruitment.

We will initially aim to recruit 90 patients per site over a three-month period, using the Trainee Emergency Research Network (TERN) and involving approximately 100 sites. We have based site numbers on expressions of national interest for participation in TERN projects, participation in previous TERN projects and the results of a recent prospective service evaluation.

Our recruitment period has been set at 3 months, using predicted attendance figures of 31 per month. Assuming a total of 100 sites actively recruiting, this will provide an expected total recruitment pool of over 9300 patients within 3 months. Should the site number be reduced, the recruitment period would extend in tandem.

## Ethics and Dissemination

### Ethical Approval

This project has ethical approval from South West Frenchay Ethics Committee (19/SW/0243) and regulatory approval from the Health Regulation Authority (UK), Health and Care Research Wales (Online Supplementary 1).

### Risk to participants

The study involves no change to clinical care and no additional interventions for participants and therefore carries no additional clinical risk. As mentioned earlier, the study design ensures patients are robustly informed of the study and are provided with information regarding their involvement and how to withdraw consent should they wish to.

### Risk to investigators

There are no anticipated additional risks to investigators as part of this study. The study may generate media interest. All media releases will be conducted through the Sponsor and/or publishing journals. Media interviews will be undertaken by a senior member of the study group with media training.

### Dissemination

The results from this study will be submitted for publication in leading journals and to national conferences for presentation. The Trainee Emergency Research Network (TERN) aims to be a promoting force for research opportunity in emergency medicine and will be providing regular input to the national academic conference agenda for emergency medicine, to highlight recent projects, ongoing work and opportunity for submission of ideas.

In addition, this project offers the opportunity for TERN to develop a national strategy with regards to project output and knowledge translation. Recent collaborations with online learning resources ensure that this project will be regularly appraised and updated throughout delivery, through a dedicated TERN section on the college website. [32]

TERN will also aim to develop a local dissemination strategy using trainees to highlight findings and implement change where relevant within local departments. Free Open Access Medical Education channels will be used to highlight and appraise the project results. Blogs, podcasts and discussions will be planned to highlight lessons learned about trainee networks research and cloud-based data collection.

## Discussion

Acute severe headache and the diagnosis of SAH remains a diagnostic problem for Clinicians. This is due to the significant mortality and morbidity associated with SAH, making it a diagnosis not to be missed. However, this must be weighed against the risks of increasing the frequency of unnecessary investigations.

The most recent meta-analysis reported CT brain sensitivity as high as 99.2% (95% CI 92.6 to 100). [33] This data includes the most recent publication in 2020 by Perry et al in which sensitivity was reported as 95.5% (95% CI 91.6-99.4%). [13] The wide confidence intervals are a concern and drop the sensitivity below the desired 99% sensitivity as expressed by EM physicians. [30] Confirmation of the sensitivity of CT brain within 6 hours in UK setting remains a research priority.

The specificity of CT brain within 6 hours, 100% (95% CI 99.0-100%), is reassuring. [33] The accurate identification of patients who are disease (SAH) negative with a negative test (CT brain), ensures patients are not unnecessarily exposed to further invasive procedures. However, the same cannot be said about the SAH clinical decision rule. Despite its promising reported sensitivity of 100% (95% CI 98.1%-100%), the specificity was 12.7% (95% 11.7% - 13.9%). There is a concern that widespread adoption of the SAH clinical decision rule could actually lead to more investigations, not less. The original authors suggest: “Patients require investigation if one or more finding is present”. [13] With such a low reported specificity this could have a significant impact on how patients with headache are managed, and wider ramifications for other patients and staff in the ED. Determining the test characteristics in the UK will allow a better understanding of performance and allow a more informed discussion about how and if it should be integrated into UK practice.

The final area this research will examine is the test characteristics of CT brain after 6 hours. The initial focus of the 6 hour interval was based on the median interval between headache onset and imaging in positive SAH cases in the original study by Perry. [9] Given that 50% of patients present outside of this timeframe and there is clear lack of biological plausibility for a dichotomous time cutpoint, there is a pressing need to determine the accuracy of CT brain beyond 6 hours. This data will aid balanced clinical assessment and facilitate shared decision making between clinicians and patients. From a UK perspective, the accuracy of CT head in diagnosing SAH at 12 and 24 hours has recently been identified as a research priority by NICE. This work will take preliminary steps towards addressing that research question. [20]

Beyond the diagnostic accuracy of imaging and decision rules, there is ongoing debate about the gold standard for diagnosis of SAH in research studies. This protocol has adopted the definition of SAH proposed by Perry et al to provide consistency and comparison with their work. However, their most recent study provides a clear example of some of the limitations of this definition. Their final criterion for diagnosis of SAH is:

*“Red blood cells (>5×10*^*6*^ */L) in the final tube of cerebrospinal fluid collected **<u>and</u>** an aneurysm identified on cerebral angiography (digital subtraction, computed tomography, or magnetic resonance angiography)”*

This resulted in two patients being classified as false positives for CT brain within 6 hours (i.e. blood identified on LP and aneurysm present on CT-A). This was despite the treating neurosurgeon in each case diagnosing a traumatic LP and incidental aneurysm (i.e true negatives). Due to very small number of false positives within the cohort, this has a significant effect on the reported sensitivity. This potential limitation within our study design, has been mitigated against with the use of the UK 2008 National Biochemist reporting guideline where possible and the use of an adjudication committee.

In conclusion this protocol outlines a robust methodology for undertaking a large observational study to report the test characteristics of early neuroimaging following acute severe headache and the test characteristics of the Ottawa SAH clinical decision rule. Our study will directly address research priorities highlighted by the James Lind Alliance Emergency Medicine Priority setting partnership and NICE in the UK. [19,20]

## Data Availability

N/A

## List of Abbreviations

(ED): Emergency Department
(UK): United Kingdom
(SAH): Subarachnoid Haemorrhage
(CT): Computerised Tomography
(NICE): National Institute for Health and Care Excellence

## Declarations

### Ethics approval and consent to participate

This project has ethical approval from South West Frenchay Ethics Committee (19/SW/0243). There is ethical approval for ‘opt-out’ consent, which means consent is assumed unless participant ‘opt-out’. This can be done verbally or in writing. This process is explained in detail in the consent section above.

### Consent for publication

Not applicable as a study protocol

### Author Contributions

TR conceived the idea for the study. TR and DH, were responsible for the initial study design, which was refined with the help of WH. WH provided the statistical plan. TR and RH will lead the dissemination of the study in UK Adult EDs. TR & RH will coordinate study set-up, finalise the study surveys. All authors contributed to the final study design and protocol development, critically revised successive drafts of the manuscript and approved the final version. The study management group is responsible for the conduct of the study.

## Acknowledgements

The views expressed are those of the authors and not necessarily those of the NHS, the NIHR, the Department of Health or the Royal College of Emergency Medicine involved in The authors would like to acknowledge Mai Baquedano at the University of Bristol, for her support with REDCap and Beverley Greenhalgh at Salford Royal NHS Foundation Trust for her work as sponsor representative during study set up.

## Availability of data and materials

Not applicable as a study protocol

